# Demographic and psychological correlates of SARS-CoV-2 vaccination intentions in a sample of Canadian families

**DOI:** 10.1101/2020.11.04.20226050

**Authors:** Christine L. Lackner, Charles H. Wang

**Author notes:** Note: Ethical clearance was obtained from the Mount Saint Vincent University Research Ethics Board, according to the TriCouncil Policy Statement: Ethical Conduct for Research Involving Humans.

## Abstract

The COVID-19 pandemic has been ongoing for close to a year, with second waves occurring presently and many viewing a vaccine as the most likely way to curb successive waves and promote herd immunity. Reaching herd immunity status likely necessitates that children, as well as their parents, receive a vaccine targeting SARS-CoV-2. In this exploratory study, we investigated the demographic, experiential, and psychological factors associated with the anticipated likelihood and speed of having children receive a SARS-CoV-2 vaccine in a sample of 455 Canadian families (857 children). Using linear mixed effects and proportional odds logistic regression models, we demonstrated that older parental age, living in the Prairies (relative to Central Canada), more complete child and parental vaccination history, more positive attitudes towards vaccines generally, higher psychological avoidance of the pandemic and a greater tendency to prioritize the risks of the disease relative to the risks of side effects (i.e., lower omission bias), were associated with higher likelihoods of intention to vaccinate participants’ children. In some models, subjective evaluations of proximal COVID-19 risk and higher levels of state anxiety were associated with increased likelihood of having children vaccinated. Faster speed of intended vaccination was predicted by a similar constellation of variables, with higher SES emerging as a trend-level predictor of vaccination speed. Results are discussed with respect to public health knowledge mobilization.

---

While a vaccine is currently unavailable for the novel coronavirus (SARS-CoV-2, the virus), several pharmaceutical companies and research groups are making progress [1–3]. However, even if a successful vaccine is developed, enough people need to be willing to receive the vaccine to achieve herd immunity status, as natural exposure may be insufficient to reach this level of protection [4]. While the proportion of vaccinated individuals needed to achieve herd immunity status varies by disease, projected estimates for COVID-19 (the disease) range between 56% and 82% [5–7], and some believe these proportions are impossible to estimate for COVID-19 [4]. Further, 19.2% of the Canadian population is aged younger than 18 years [8], which likely necessitates that children be vaccinated to achieve these values, considering refusal rates in other segments of the population (e.g., only 30.8% of Canadians aged 18-64 without a chronic medical condition were vaccinated against influenza in the 2018-2019 season, [9]).

Vaccine hesitancy and anti-vaccine convictions pose a significant public health challenge globally [10]. Even in Canada, where vaccines are free to administer and relatively easy to access, many parents refuse to completely or partially vaccinate their children—both for regularly scheduled vaccines (e.g., measles, mumps, and rubella [MMR]; pertussis) and seasonal vaccines (e.g., influenza). Canada has failed to meet its goal of 95% vaccine coverage in accordance with the World Health Organization [11] and ranks 28th out of 29 affluent countries for vaccine coverage rates [12]. Therefore, studying intentions to vaccinate against SARS-CoV-2 *in the Canadian context* is especially important if Canada wants to advance its ranking and promote herd immunity against COVID-19 while recognizing a degree of generalizability of our findings to other similar countries.

In addition to the notion that children will need to be vaccinated against COVID-19 to reach herd immunity status within both the larger community and subpopulations with which they regularly interact (e.g., childcare and school peers), it is important to study vaccination intentions *in the family context* for the following reasons. (1) parents are the health decision makers for their children; (2) children, relative to adults, are less likely to be COVID-19 symptomatic, and, in turn, less likely to fully isolate [13], which may increase the likelihood that they will transmit the disease to others (although see ref [14]). Thus, understanding parental intentions to vaccinate themselves and their children for SARS-CoV-2 is essential. Families are a pivotal piece of the public health landscape. In this exploratory study, we examined what demographic, experiential, and psychological factors predict Canadian parents’ intentions to have their family vaccinated.

## Demographic predictors

There are numerous demographic predictors of vaccination intentions and behaviours. Increased family size is negatively correlated with likelihood of being immunized against influenza [15], pertussis [16], [17], and vaccine completeness at 19 months of age ([18], although see ref [19]). Those of higher socio-economic status (SES), more advanced age, and from non-minority backgrounds are more likely to intend to vaccinate, or actually receive the vaccination for seasonal influenza, H1N1, and/or have their children vaccinated against MMR [15], [20]– [24]. Adolescents in rural areas of the USA (vs. urban) are less likely to initiate and complete human papillomavirus (HPV) vaccination [25], [26]. Individuals living in impoverished areas are less likely to be vaccinated against influenza [15], [27] and H1N1 [23] than individuals in affluent areas. Further, indicators of vaccine doubt differ across regions of the USA, with those in the West more likely to refuse or delay their children’s routine vaccinations than those in other census areas [28]. Thus, we explored the demographic predictors of likelihood and speed of having children receive a SARS-CoV-2 vaccination to see if previous relationships hold in the novel public health context of COVID-19 or if they are washed out by psychological and experiential predictors that may be specific to COVID-19.

## Psychological and experiential predictors

Given that vaccine uptake is not uniform across populations (i.e., there are individual difference variables to be considered; [28], we explored the psychological and experiential predictors of intentions to vaccinate against SARS-CoV-2. We examined psychological and experiential predictors that are both specific to COVID-19 (e.g., knowing someone with the disease) and non-specific to COVID-19 (e.g., levels of trait anxiety).

First, we examined previous experience with vaccination: the completeness of vaccine history and the experience of vaccine adverse events (VAEs) as potential predictors of both the speed and likelihood of having children vaccinated. Individuals who regularly vaccinated against influenza were more likely to be vaccinated against H1N1 during the 2009 epidemic [23], [29– 31]. Mothers who decline influenza vaccination during pregnancy are less likely to have their children fully vaccinated [32]. Vaccine history is associated with MMR vaccine uptake [33]. Therefore, those with complete vaccination schedules may be more likely to have their children receive the SARS-CoV-2 vaccine than those with incomplete schedules.

Child vaccination history is predicted by parental attitudes towards vaccines and the experience of, or worries about, VAE. Concerns over VAEs influence parental attitudes towards immunization beginning when their child is an infant [34]. Early experiences with vaccines can have enduring influences on subsequent vaccinations. Parents who have personal experience with or knowledge of others with VAEs have lower confidence ratings in the safety, health benefits, and effectiveness of vaccination [35], and they have higher levels of vaccine hesitancy than those with less experience or knowledge [36]. Exposure to vignettes involving VAEs decreases participants’ intentions to get vaccinated against a hypothetical disease [37]. Clearly, VAEs play a significant role in parental attitudes and behaviours regarding vaccines (e.g., ref [28]). Thus, we asked parents about their own, and their children’s, experiences with VAEs (if any). Additionally, attitudes towards vaccinations generally were assessed, including their perceived dangers, powerlessness, and trust in authorities regarding vaccines—which all influence vaccination intentions [19], [38], [39].

We also asked participants about whether they and their children had a primary care physician. Having a strong, trusting relationship with a primary healthcare provider is associated with increased confidence in vaccines [35]. Doctor’s recommendations for vaccination were associated with increased H1N1 vaccine uptake [31]. We wanted to know whether the same pattern would hold true for SARS-CoV-2 vaccination intentions.

Experiential variables such as vaccine history, VAEs, and access to care cannot likely be separated from the psychology of individuals and their thinking about vaccination. Additional psychological variables may also influence opinions about vaccination. First, arguably, the closer the disease “hits home” the greater the likelihood of taking action to protect oneself and one’s family. For instance, compared to their counterparts, young women with a family history of gynecological cancers are more likely to get the HPV vaccine [40], and children with a family history of immigration from a highly tuberculosis endemic country are more likely to be vaccinated for tuberculosis [41]. Qualitative reports suggest that individuals with a family history of autism (vs. not) are less likely to have their children vaccinated against MMR [42], a vaccine that has been falsely pointed to for increasing risk of autism. People who believe that they had a low risk of H1N1 infection were not likely to get vaccinated against H1N1; however, they believed that their attitudes would change if a member of their social circle contracted the illness [24]. These lines of evidence converge to suggest that the socially closer the perceived threat, the greater the tendency to vaccinate.

White and colleagues describe that social distance is one component of a greater construct known as psychological distance, which influences the probability of perceiving and reacting to disease threats [43]. Therefore, we investigated how many (if any) people the responding parent knows who have been diagnosed with COVID-19, their relationship closeness, and their health outcome. The threat of COVID-19 may seem greater for those individuals with high numbers of contacts diagnosed and/or knowledge of those with serious COVID-19-related outcomes (e.g., death of a close contact leading to decreased psychological distance from COVID-19). A Malaysian study confirms this hypothesis. Knowing a friend, neighbour or colleague infected with COVID-19 was associated with greater intention to vaccinate against SARS-CoV-2 [44], although we do not know if this relationship holds when making vaccination decisions for one’s children, or in the Canadian context where vaccines are freely available. Relatedly, the perceived risk of acquiring the disease impacts vaccination decisions [45], including decisions around oneself receiving the SARS-CoV-2 vaccine [44], [46], [47] and likely impacts how psychologically distant one feels from COVID-19. To our knowledge, no one has investigated how distal COVID-19 threat (e.g., risk to community and the world) impacts SARS-CoV-2 vaccination intentions in the family context.

Perhaps related to previous experience with VAEs, omission bias—or having a perceived greater risk of harm from being vaccinated (i.e., side effects) relative to the perceived risk of not being vaccinated (i.e., riskiness of the disease)—leads to a tendency to prefer inactive options (i.e., not being vaccinated, [48]). Thus, we examined parental levels of the omission bias following Hamilton-West [48].

We additionally examined the influence of the pandemic on parents’ psychological well-being, hypothesizing that those who were most negatively affected by the pandemic would be those who were more likely to vaccinate their children as compared to their less affected counterparts. A further psychological variable, levels of state and trait anxiety may additionally correlate with intentions to vaccinate as mothers high in trait anxiety are *less likely* to have completely vaccinated their children than mothers low in anxiety [49]. Healthcare workers who believed the H1N1 vaccine was unsafe were higher in state anxiety than those who felt it was safe [50]. However, mothers with mild anxiety symptoms are *more likely* to receive the influenza vaccine during pregnancy than women without such anxiety symptoms [51]. These discordant results may be attributable to making healthcare decisions for oneself versus one’s children, measures of state versus trait anxiety, and/or the nature of the vaccine and disease in question. Levels of state anxiety may be elevated in the current pandemic context (e.g., [52–54]) as parents report stressors related to relationships, health, safety, work, and finances [55] and this increased stress may be associated with increased *or* decreased intentions to vaccinate. These lines of research suggest that it is prudent to examine associations between state and trait anxiety and intentions to vaccinate children.

Relative to previous research, we expect the pattern of those receiving vaccines to shift somewhat considering the current COVID-19 pandemic. Previous research revealed that vaccine uptake is not uniform across vaccines (e.g., rates of regularly scheduled immunizations and seasonal influenza vaccination are not equivalent, and intentions to vaccinate against H1N1 were higher than intentions to vaccinate against seasonal flu; [28], [56]), and that parental attitudes differ across vaccines (e.g., the high degree of concern over the varicella vaccine in ref [28]). Thus, in the present investigation all associations were tested with two-tailed statistical tests. Importantly, we cannot entirely look to existing vaccine literature to predict vaccination intentions during the current COVID-19 crisis; therefore, the present study, while exploratory in nature, makes a valuable contribution to our understanding of which Canadian families are likely to have their children vaccinated and how quickly.

Thus, our objectives were to determine the demographic, experiential, and psychological predictors of intent to vaccinate children for SARS-CoV-2. We examined predictors of immunization intentions including demographic variables: parental age, income, education level, health status, and family size/composition. Additionally, we examined whether the impact of the pandemic on parental mental health, social distance from COVID-19, individual differences in anxiety, attitudes towards immunizations, previous experience with VAEs, and access to primary care physicians predict future vaccination intentions for parents and their children.

## Methods

### Participants

A total of 673 participants clicked on the survey link. Forty-one participants did not advance past the consent form and 45 participants did not advance past the eligibility survey, requiring them to indicate that they were a Canadian parent of (a) child(ren) age younger than 18 years of age. Further, 132 participants failed to complete at least 80% of the survey measures and were therefore excluded from the final sample. A total of 455 parents responded to 80–100% of our measures and were therefore retained in our sample, reporting on a total of 857 children. Given that participants dropped out at varying time points throughout the survey (many before demographics were completed), we cannot systematically compare those that were retained for analysis versus those who were excluded. Demographic characteristics of the final sample and other descriptive statistics are presented in Table 1.

**Table 1.** Demographic and descriptive statistics of sample.

| Variable | N(%) | Mean | SD | Possible range |
| --- | --- | --- | --- | --- |
| Responding parent age |  | 38.2 | 6.82 | 18 and up |
| Responding parent sex |  |  |  |  |
| Female | 418(91.9) |  |  |  |
| Male | 33(7.3) |  |  |  |
| Relationship to child(ren)<sup>1</sup> |  |  |  |  |
| Biological mother/father | 801(93.5) |  |  |  |
| Legal guardian or adoptive parent | 29(3.4) |  |  |  |
| Step mother/father | 21(2.5) |  |  |  |
| Foster parent | 3(.3) |  |  |  |
| Grandmother/father | 2(.2) |  |  |  |
| Region of residence |  |  |  |  |
| Central | 187(41.1) |  |  |  |
| Atlantic | 177(38.8) |  |  |  |
| Prairies | 57(12.5) |  |  |  |
| Western | 33(7.3) |  |  |  |
| Marital status |  |  |  |  |
| Married or cohabiting | 404(88.8) |  |  |  |
| Single | 51(11.2) |  |  |  |
| Number of children |  | 1.89 | .83 | 1–6 |
| Socio-economic status (z-scored) |  | -.008 | .80 | -2.98–1.39<sup>2</sup> |
| Health variables<sup>3</sup> |  |  |  |  |
| Proportion of family members with doctor |  | .94 | .18 | 0–1 |
| Child(ren)'s number of COVID-19 health risks |  | .19 | .44 | 0–3<sup>2</sup> |
| Child's past vaccine completeness |  | 6.38 | 1.71 | 2–8 |
| Child previous VAE |  |  |  |  |
| No | 782(91.2) |  |  |  |
| Yes | 75(8.8) |  |  |  |
| Parent's past vaccine completeness |  | 6.0 | 1.32 | 2–8 |
| Parent's previous VAE |  |  |  |  |
| No | 403(88.6) |  |  |  |
| Yes | 46(10.1) |  |  |  |
| Parent's number of COVID-19 health risks |  | .93 | 1.15 | 0–8<sup>2</sup> |
| Attitudes towards vaccines |  |  |  |  |
| Perceived danger |  | 21.48 | 11.84 | 8–56 |
| Powerless |  | 8.21 | 4.41 | 3–21 |
| Trust in authority |  | 7.87 | 2.98 | 2–12 |
| Omission bias |  | -3.56 | 10.18 | -49–23 |
| COVID-19 risk perceptions |  |  |  |  |
| Proximal COVID-19 risk |  | 6.18 | 2.03 | 1–10 |
| Distal COVID-19 risk |  | 7.79 | 1.94 | 1–10 |
| Psychological distance |  |  |  |  |
| COVID-19 relationship score |  | 2.55 | 8.10 | 0–150<sup>2</sup> |
| COVID-19 outcome score |  | 1.83 | 5.02 | 0–60<sup>2</sup> |
| Impact of COVID-19 pandemic event |  |  |  |  |
| Intrusion |  | 17.17 | 5.74 | 8–32 |
| Avoidance |  | 18.04 | 4.83 | 8–32 |
| Arousal |  | 12.47 | 4.42 | 6–24 |
| Anxiety |  |  |  |  |
| State |  | 43.48 | 12.61 | 20–80 |
| Trait |  | 42.17 | 9.70 | 20–80 |
| Outcome variables |  |  |  |  |
| Average child vaccination likelihood |  | 76.83 | 33.32 | 1–100 |
| Average child vaccination speed<sup>4</sup> |  | 2.46 | 1.39 | 1–5 |

Notes: <sup>1</sup>relationship could differ across children. For instance, the responding parent may be a biological parent to one child and a foster parent to another. Therefore, these statistics are presented at the child level, total $n = 857$ .
<sup>2</sup> indicates observed range, as z-scores or health conditions could theoretically take any value
<sup>3</sup> averaged at the family level for descriptive purposes
<sup>4</sup> higher values indicate a slower intended speed

### Procedure

Participants were recruited to participate in “A study of health behaviours and intentions in the wake of COVID-19” from across Canada using online advertisements and snowball sampling techniques. To avoid biased sampling, we did not include specific reference to vaccinations in our recruitment strategies. A recruitment notice was posted to the Kijiji websites of major Canadian cities, various Canadian parenting groups on Facebook, and to a variety of academic listserves. Targeted Facebook ads were visible to Canadian parents aged 18– 60 years for a two-week period during data collection, which lasted from May 15 to June 9, 2020. The recruitment notices contained a link to SimpleSurvey^™^ —where participants read an informed consent letter and indicated their willingness to participate. Questionnaire completion took approximately 30 minutes. Both the informed consent letter and the thank-you message contained links to mental health and COVID-19 resources. Ten $50 gift cards were raffled off to randomly selected participants after all data were collected. All aspects of the study were approved by the Mount Saint Vincent University Research Ethics Board (File 2019-197), abiding by the principles of the Tri-Council Policy Statement: Ethical Conduct for Research Involving Humans.

### Materials

#### Family variables

We asked how many children aged younger than 18 years are currently under their care, how many adults aged 18 years or older live in the home, and how many adults aged 65 years or older (an at-risk demographic) live in the home. Parents were asked to report the children’s ages and their relationship to the child (e.g., biological mother/father, stepmother/father, etc).

#### Demographics questionnaire

We asked parents to report on their age, sex, province of residence, residential population density (urban, rural, mixed), work status, occupation, family income, marital status, and highest level of education completed. Province of residence was categorized into Atlantic (Nova Scotia, New Brunswick, PEI, and Newfoundland), Central (Quebec and Ontario), Prairies (Manitoba, Alberta, and Saskatchewan), and Western (British Columbia). If participants indicated that they were married, we asked them to report on their spouse/partner’s work status, occupation, and highest level of education completed. Family size, annual family income, and residential population density were used to calculate income-to-needs ratios per the 2018 low-income cut-offs [57] Parental occupation was coded according to the 9-level Hollingshead Index [58] by a trained research assistant. If two parents lived in the home, averaged Hollingshead scores and educational levels were calculated. Averaged Hollingshead scores, averaged educational levels, and income-to-needs ratios were *z*-scored and then summed to form a measure of SES.

### Health history

#### Access to a primary care physician

The responding parent was asked to report, for each child and their spouse/partner (if applicable), if they have a family doctor or pediatrician. From this we calculated the proportion of family members with a doctor. All child-level variables were responded to from oldest child to youngest child and therefore birth order was coded for.

#### Pre-existing risk factors for COVID-19

The responding parent was asked to select from a list of 17 cardiac, respiratory, weight-related, and immune system conditions (e.g., congestive heart failure, chronic lung disease, obesity, HIV, etc.), which, if any, they had been diagnosed with. They were given the option to provide other diagnoses which were subsequently coded as a risk factor for COVID-19 or not. They completed this same checklist for each child. From this checklist, we summed the number of COVID-19-related health risks the responding parent and each child possessed. Parental COVID-19 health risks were treated as a continuous variable (Range 0 - 8), while child COVID-19 health risks were dichotomized to 0, 1, or 2+ risks given the relatively fewer health risks reported in this group.

#### COVID-19 health status

One item was used to assess the responding parents’ current COVID-19 related health status. They were asked over the last two weeks if they had been perfectly healthy, had been ill but do not believe it is COVID-19, had been ill and know it is not COVID-19 because they tested negative, had been ill and think it is COVID-19, or had been ill and know it is COVID-19 because of a positive test result. These responses were coded from one to five, respectively. These items were used, with permission, from unpublished data by Santistevan and colleagues [59], but were not included in the present analysis as few participants reported a positive (*n* = 0) or possible (*n* = 9) COVID-19 diagnosis.

#### Responding parent vaccination history

We provided parents with a list of the recommended childhood vaccines and asked if they had received all of those vaccines in at the recommended ages, all according to a delayed schedule, were partially vaccinated or not at all (scored from 1 to 4). We opted not to ask for this information on a vaccine-by-vaccine basis, as we anticipated significant memory difficulties at this single-vaccine level (e.g., some of the immunizations are administered as a baby, and we did not expect participants to remember if they received such early vaccinations in an on-time or delayed manner). We asked how frequently they get the flu shot (with options ranging from *every year* to *never*), and responses were scored from 1 to 4. Women were asked if they received the HPV vaccine (scored 0 or 1). All participants over the age of 50 years were asked if they received the shingles vaccine (scored 0 or 1). Total vaccination completeness scores were converted to a score out of 8 to account for age and sex differences in the total number of recommended vaccines.

#### Children’s vaccination history

We asked parents to complete the above vaccine completeness measures for each of their children, excluding questions about HPV and shingles vaccines.

Vaccine completeness was scored out of 8.

#### Adverse immunization reactions

We asked parents if they or each of their children had ever experienced an adverse reaction to immunization. This was coded dichotomously (yes/no). Psychological variables.

#### Attitudes towards vaccines

We assessed participants’ attitudes towards vaccinations generally using measures from Jolley and Douglas [39]. Eight questions measured the perceived dangers of vaccines (e.g., “Vaccines lead to allergies”). Three items measured feelings of powerlessness surrounding vaccination (e.g., “I feel that my actions will not stop the negative outcomes of immunizations”). Perceived dangers and powerlessness were measured on a Likert scale ranging from 1 (*strongly disagree*) to 7 (*strongly agree*), and subscale scores were calculated by summing responses. Two questions measured trust in authorities (e.g., “I trust corporations to tell the truth about vaccinations”) on a Likert scale ranging from 1 (*strongly distrust*) to 6 (*strongly trust*). Responses to these two items were summed. Jolley and Douglas reported good reliability of these four measures (all αs ≥ .82).

#### Omission bias

The strength of the omission bias concerning the COVID-19 vaccine was measured using modified materials from Hamilton-West (2006) [48]. All references to the MMR vaccine were replaced with the SARS-CoV-2 vaccine, and the questionnaire was prefaced with the following: “Various research and pharmaceutical companies are in the process of developing a vaccine against SARS-CoV-2 (the virus) to reduce the risk of developing COVID-19 (the disease). Thinking towards this future possibility, for each of the questions below, choose the response that best characterizes your attitudes.” To calculate the omission bias total, two variables were needed: (1) risks associated with not vaccinating (total COVID-19 risk perception mean x risks of COVID-19 if not vaccinated) and (2) risks associated with vaccinating (risk of COVID-19 if vaccinated x total COVID-19 risk perception, plus risk of COVID-19 vaccine side effects × severity of side effects). The risks associated with vaccinating were subtracted from the risks associated with not vaccinating, and negative scores indicate perceived greater risk associated with vaccinating, and a likely tendency to prefer inaction/non-vaccination (i.e., negatives scores are associated with greater omission bias).

#### COVID-19 risk perceptions

Participants were asked to rate, on a scale from 1 (*no risk*) to 5 (*very serious risk*), how much risk COVID-19 poses to them personally, to their family, to their community, and to humankind (4 items). Personal and family ratings were summed (proximal risk), as were community and humankind ratings (distal risk). These items were used, with permission, from unpublished data by ref [59].

#### Psychological Distance

To measure psychological distance, we asked participants how many people that they knew who had been diagnosed with COVID-19 and what their relationship was to that person(s) (ranging from an immediate family member living in the same household (scored a 6) to a friend or relative of one of your acquaintances (scored a 1)). The number of positive diagnoses per relationship category were summed in a weighted fashion to yield a COVID-19 relationship closeness score. Further, participants were asked to report on the individual(s)’ health outcomes ranging from “they are currently recovering at home and did not require hospitalization” (scored a 1) to “they passed away” (scored a 5). The health outcomes were multiplied by the number of people in that outcome category to yield a COVID-19 outcome score.

#### Impact of the COVID-19 pandemic event

The subjective impact of the COVID-19 pandemic and its associated restrictions was assessed using the Impact of Event Scale-Revised [60], [61]. The questionnaire was modified only to include the preface that participants should respond concerning how the COVID-19 pandemic has made them feel. The Impact of Event scale comprised 22 questions forming 3 subscales: the Intrusion subscale includes 8 items (e.g., “Any reminders brought back feelings about it”), the Avoidance subscale includes 8 items (e.g., “I avoided letting myself get upset”), and the Arousal subscale includes 6 items (e.g., “I felt irritable and angry”). All responses were scored on a Likert scale ranging from 1 (*not at all*) to 4 (*often*).

#### Anxiety

The State-Trait Anxiety Inventory for Adults (Mind Garden Inc., CA) was used to evaluate parental levels of state and trait anxiety. State anxiety was measured using 20 Likert-style questions, evaluated on a scale from 1 (*not at all*) to 4 (*very much so*), and participants responded to items such as “I feel at ease” and “I feel upset” concerning how they are feeling at the present moment. An additional 20 items measured trait anxiety. Participants were asked to respond to items such as “I lack self-confidence” and “I am a steady person” on a scale from 1 (*almost never*) to 4 (*always*), concerning how they generally feel, excluding their feelings during the COVID-19 pandemic to the best of their ability. Scores on the state and trait subscales could range from 20 to 80, with higher scores indicating higher levels of anxiety. Outcome variables.

#### SARS-CoV-2 vaccination intentions

Participants were asked to report, on a scale from 1 to 100, how likely they would be to receive a vaccination and how likely they would be to have their children vaccinated against SARS-CoV-2 in the event that a successful vaccine is developed and approved by Health Canada. They were also asked how quickly they would get themselves and each of their children vaccinated ranging from 1 (*as soon as the vaccine is available in my area*) to 5 (*I would not get them vaccinated for SARS-CoV-2*). They responded to these likelihood and speed questions for each of their children.

### Data analysis

All data were processed and analysed using R (Vienna, Austria). To model the likelihood of parents intending to have their child(ren) vaccinated, we first fit a multiple linear regression model with the independent variables (IVs), shown in Table 1 (all except the variable “Relationship to child(ren)” and the two outcome variables). Parents reported on the likelihood of vaccinating each of their children, and each child is an observation in the model fitting. In this analysis, child birth order was one categorical IV with any third or subsequent children were combined to form a birth order category of 3+ (having 4, 5, or 6 children was relatively rare). Variables specific to an individual child (e.g., their age, vaccine history) were used to model that child’s vaccine likelihood only, whereas parental or family level variables (e.g., family size, SES, Canadian region of residence, pandemic-related variables), were used to model all children’s vaccine likelihood within a family.

The above multiple regression model did not take into account the natural dependencies among the observations, as children were nested within families, which in turn were nested within Canadian region of residence. It is reasonable to think parents tend to take similar actions on vaccination toward each of their children and that there may be similar trends in vaccination intention in the same geographic region. Multi-level Linear Mixed-effects models [62] were therefore considered. Models included random intercepts allowing for children nested within their family and families nested within the same geographic region. The lme4 method [63] in R was used to run a multi-level mixed model with random intercepts.

Multicollinearity among the IVs in the linear and linear mixed-effects models was addressed using a variance inflation factor (VIF) analysis. VIF greater than 5 indicates significant multicollinearity and the corresponding variable may need to be removed from the models. All variables in the linear model had a VIF of less than 3.81 and therefore multicollinearity was not a significant problem. For the multi-level mixed model with random intercepts, the “baby” group (aged less than 24 months) had a marginally high VIF at 5.5, but we retained this variable to avoid losing data for this group of children and the families to which they belong.

To analyse the speed of SARS-CoV-2 vaccination, a proportional odds logistic regression (polr) model for ordinal logistic regression [64] was applied. Under this model, the odds of being less than or equal a particular category (e.g., the lowest category “as soon as the vaccine is available” to all categories above it) is the same as the odds being the two lowest categories “as soon as the vaccine is available” and “a couple of weeks after the vaccine is available” to the other categories above them, etc. The cut points between the categories are the estimated intercepts (there are four of them resulting from five categories of vaccination speed). The estimated slope coefficients are the log odds ratio associated with one-unit change in the corresponding predictor variables. The exponential of the log odds ratio is therefore the odds ratio of an outcome being Y > a certain category (say, *j*) to the outcome Y ≤ *j* when the corresponding predictor (x) changes by one unit. If the odds ratio is greater (or less) than 1, then the odds of the outcome Y > j increases (or decreases) with the variable x by the amount of the odds ratio.

Vaccination speeds are not independent from each other, as there also exists a nested relationship among children, family, and region of residence. However, the polr model implemented in R cannot include random-effects terms. We therefore tried Cumulative Link Mixed Models (clmm) implemented in the “ordinal” package [65] to conduct an ordinal regression with random intercepts.

## Results

### SARS-CoV-2 vaccination likelihood

The results of the highly significant linear regression model can be seen in Table 2. The model captured 65% of the variation in the outcome variable – the likelihood of vaccination. Participants in the Prairies (relative to the Central provinces) reported a higher likelihood of having their children vaccinated. No other regional differences were significant. Older parental age was associated with increased likelihood. At the child level, only vaccine history was a significant independent predictor of vaccination likelihood and children who had higher vaccine completeness showed higher likelihood to get vaccinated for COVID-19. At the family level, more complete parental vaccination history, lower perceived vaccine danger, higher trust in authority, higher omission bias, higher proximal perceived COVID-19 risk, higher levels of avoidance, and higher levels of state anxiety were all significant predictors of increased child vaccination likelihood.

**Table 2.** Linear regression model of the likelihood of having children vaccinated (multiple R^2^ = 0.65)

|  | Estimate | Std. Error | t value | Pr(>\|t\|) |
| --- | --- | --- | --- | --- |
| Intercept | 12.16 | 12.85 | 0.9 | .34 |
| Demographics |  |  |  |  |
| Parent sex – Male (female as ref grp) | -4.11 | 3.13 | 1.3 | .19 |
| Parent age | 0.47 | 0.17 | 2.8 | .005 |
| Family size | -0.40 | 0.99 | 0.4 | .69 |
| SES | -1.21 | 1.14 | 1.1 | .29 |
| Child age – baby as ref grp |  |  |  |  |
| Preschool | -1.39 | 2.46 | 0.6 | .57 |
| Child | -1.18 | 2.69 | 0.4 | .66 |
| Adolescent | 0.93 | 3.55 | 0.3 | .79 |
| Birth order – first born as ref grp |  |  |  |  |
| Second born | -1.42 | 1.78 | 0.8 | .43 |
| Third or later born | -0.87 | 3.24 | 0.3 | .79 |
| Region of residence – Central as ref grp |  |  |  |  |
| Atlantic | 2.71 | 1.79 | 1.5 | .13 |
| Prairies | 8.29 | 2.51 | 3.3 | .001 |
| Western | 2.57 | 3.16 | 0.8 | .42 |
| Health variables |  |  |  |  |
| Proportion of family members with doctor | 4.1 | 4.73 | 0.9 | .39 |
| Child has 1 COVID-19 health risk (ref grp = 0 risks) | 2.96 | 2.45 | 1.2 | .23 |
| Child has 2+ COVID-19 health risks (ref grp = 0 risks) | 2.36 | 4.23 | 0.6 | .58 |
| Child's past vaccine completeness | 1.90 | 0.76 | 2.5 | .013 |
| Child's previous VAE (ref grp = no VAEs) | -1.01 | 2.98 | 0.3 | .74 |
| Parent's past vaccine completeness | 2.80 | 1.02 | 2.8 | .006 |
| Parent's previous VAE (ref grp = no VAEs) | <-0.01 | 2.84 | <0.001 | >.99 |
| Parent's number of COVID-19 health risks (continuous) | 0.02 | 0.77 | <0.001 | .98 |
| Attitudes towards vaccines |  |  |  |  |
| Perceived danger | -0.25 | 0.12 | 2.1 | .032 |
| Powerless | 0.04 | 0.25 | 0.2 | .87 |
| Trust in authority | 1.33 | 0.40 | 3.3 | .001 |
| Omission bias | 1.77 | 0.14 | 12.9 | <.001 |
| COVID-19 risk perceptions |  |  |  |  |
| Proximal COVID-19 risk | -1.97 | 1.04 | 1.9 | .06 |
| Distal COVID-19 risk | 1.36 | 1.21 | 1.1 | .27 |
| Psychological distance |  |  |  |  |
| COVID-19 relationship score | 0.036 | 0.13 | 0.3 | .80 |
| COVID-19 outcome score | <0.01 | 0.20 | 0.0 | .99 |
| Impact of COVID-19 pandemic event |  |  |  |  |
| Intrusion | -0.07 | 0.26 | 0.3 | .77 |
| Avoidance | 0.65 | 0.21 | 3.1 | .002 |
| Arousal | -0.41 | 0.34 | 1.2 | .23 |
| Anxiety |  |  |  |  |
| State | 0.17 | 0.09 | 2.0 | .05 |
| Trait | 0.07 | 0.11 | 0.6 | .52 |

Notes: (1) Ref grp = reference group. All categorical variables must be compared against a reference group as indicated in the table. (2) Negative scores indicate higher omission bias, so here, lower omission bias is associated with increased likelihood of having child vaccinated.

Under the multiple-level linear mixed-effects model, where the interdependence between the observations for the children in the same families and the families in the same geographic regions were taken into account, the results also showed increased parental age, more complete parent and child vaccination history, lower perceived danger, and lower omission bias were significant for predicting increased child vaccine likelihood (see Table 3). However, in this nested model, trust in authority and avoidance fell somewhat below statistical significance, but still showed trend-level predictions in the same direction as the non-nested model. Proximal COVID-19 risk was no longer associated with reduction in vaccine likelihood. An additional trend-level predictor of child age emerged, with parents reporting higher average vaccine likelihoods for adolescent children relative to babies and other young children. Levels of state anxiety were no longer significantly predictive as they were in the non-nested model.

**Table 3.**
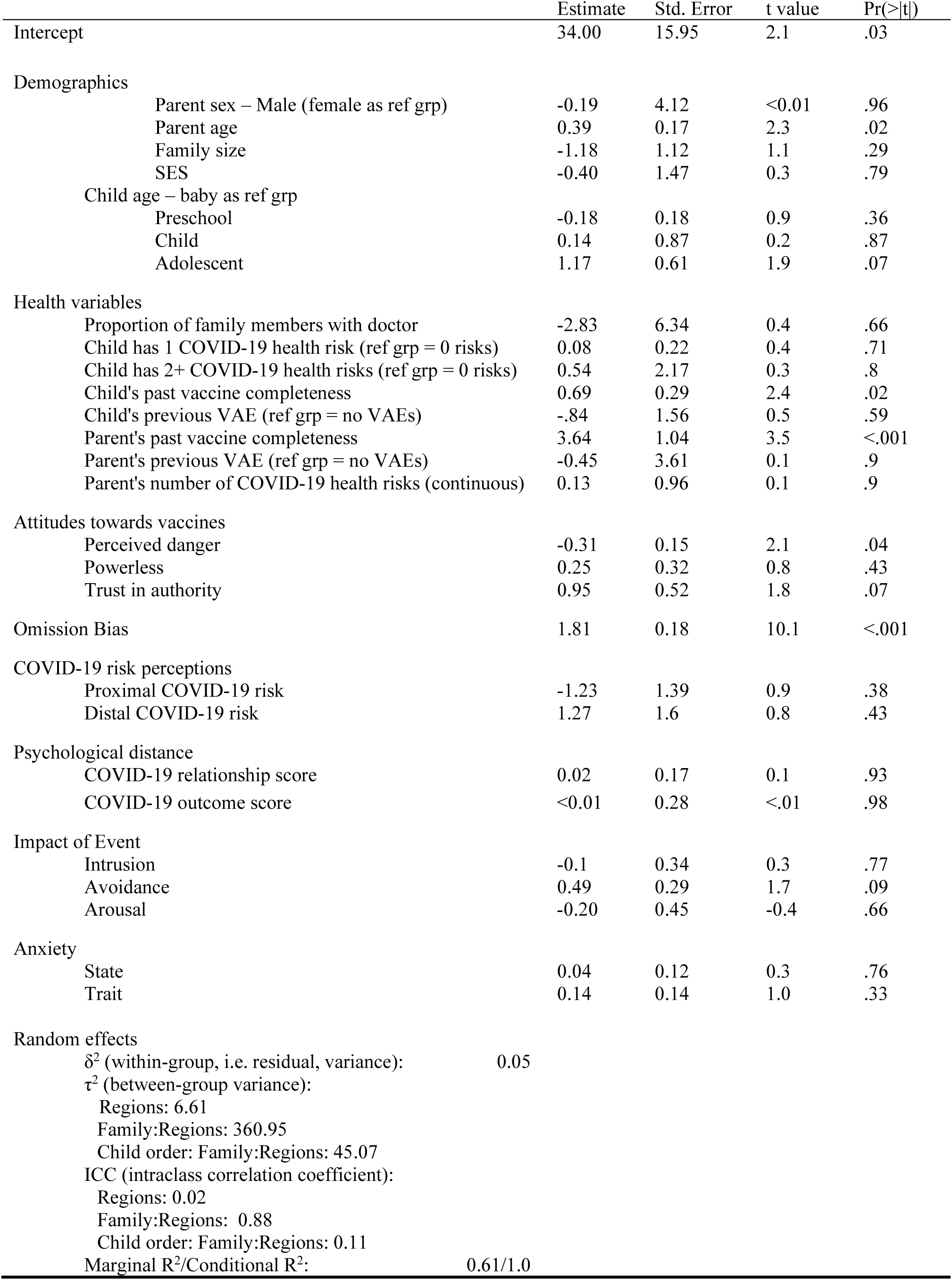
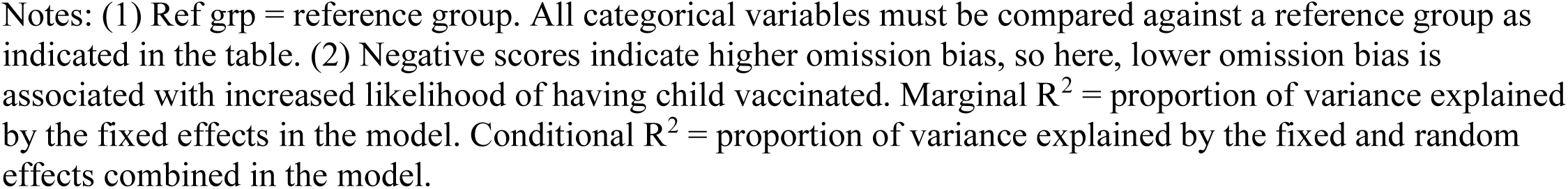
Multilevel mixed-effects regression model with random intercepts predicting likelihood of having children vaccinated

|  | Estimate | Std. Error | t value | Pr(>\|t\|) |
| --- | --- | --- | --- | --- |
| Intercept | 34.00 | 15.95 | 2.1 | .03 |
| Demographics |  |  |  |  |
| Parent sex – Male (female as ref grp) | -0.19 | 4.12 | <0.01 | .96 |
| Parent age | 0.39 | 0.17 | 2.3 | .02 |
| Family size | -1.18 | 1.12 | 1.1 | .29 |
| SES | -0.40 | 1.47 | 0.3 | .79 |
| Child age – baby as ref grp |  |  |  |  |
| Preschool | -0.18 | 0.18 | 0.9 | .36 |
| Child | 0.14 | 0.87 | 0.2 | .87 |
| Adolescent | 1.17 | 0.61 | 1.9 | .07 |
| Health variables |  |  |  |  |
| Proportion of family members with doctor | -2.83 | 6.34 | 0.4 | .66 |
| Child has 1 COVID-19 health risk (ref grp = 0 risks) | 0.08 | 0.22 | 0.4 | .71 |
| Child has 2+ COVID-19 health risks (ref grp = 0 risks) | 0.54 | 2.17 | 0.3 | .8 |
| Child's past vaccine completeness | 0.69 | 0.29 | 2.4 | .02 |
| Child's previous VAE (ref grp = no VAEs) | -.84 | 1.56 | 0.5 | .59 |
| Parent's past vaccine completeness | 3.64 | 1.04 | 3.5 | <.001 |
| Parent's previous VAE (ref grp = no VAEs) | -0.45 | 3.61 | 0.1 | .9 |
| Parent's number of COVID-19 health risks (continuous) | 0.13 | 0.96 | 0.1 | .9 |
| Attitudes towards vaccines |  |  |  |  |
| Perceived danger | -0.31 | 0.15 | 2.1 | .04 |
| Powerless | 0.25 | 0.32 | 0.8 | .43 |
| Trust in authority | 0.95 | 0.52 | 1.8 | .07 |
| Omission Bias | 1.81 | 0.18 | 10.1 | <.001 |
| COVID-19 risk perceptions |  |  |  |  |
| Proximal COVID-19 risk | -1.23 | 1.39 | 0.9 | .38 |
| Distal COVID-19 risk | 1.27 | 1.6 | 0.8 | .43 |
| Psychological distance |  |  |  |  |
| COVID-19 relationship score | 0.02 | 0.17 | 0.1 | .93 |
| COVID-19 outcome score | <0.01 | 0.28 | <.01 | .98 |
| Impact of Event |  |  |  |  |
| Intrusion | -0.1 | 0.34 | 0.3 | .77 |
| Avoidance | 0.49 | 0.29 | 1.7 | .09 |
| Arousal | -0.20 | 0.45 | -0.4 | .66 |
| Anxiety |  |  |  |  |
| State | 0.04 | 0.12 | 0.3 | .76 |
| Trait | 0.14 | 0.14 | 1.0 | .33 |
| Random effects |  |  |  |  |
| $\delta^2$ (within-group, i.e. residual, variance): | 0.05 | | | |
| $\tau^2$ (between-group variance): | | | | |
| Regions: 6.61 |  |  |  |  |
| Family:Regions: 360.95 |  |  |  |  |
| Child order: Family:Regions: 45.07 |  |  |  |  |
| ICC (intraclass correlation coefficient): |  |  |  |  |
| Regions: 0.02 |  |  |  |  |
| Family:Regions: 0.88 |  |  |  |  |
| Child order: Family:Regions: 0.11 |  |  |  |  |
| Marginal R<sup>2</sup>/Conditional R<sup>2</sup>: | 0.61/1.0 |  |  |  |

Notes: (1) Ref grp = reference group. All categorical variables must be compared against a reference group as indicated in the table. (2) Negative scores indicate higher omission bias, so here, lower omission bias is associated with increased likelihood of having child vaccinated. Marginal $R^2$ = proportion of variance explained by the fixed effects in the model. Conditional $R^2$ = proportion of variance explained by the fixed and random effects combined in the model.

### SARS-CoV-2 vaccination speed

Table 4 presents the results of a proportional odds logistic regression (polr) predicting speed of SARS-CoV-2 vaccination in children. Older parental age, living in the Prairies (relative to Central Canada), a more complete parent and child vaccine history, lower perceived danger, higher trust in authority, distal COVID-19 risk, lower omission bias, and higher levels of state anxiety, were associated with faster intended speed of vaccination. Higher SES was a trend-level predictor of faster vaccination speed. Proximal COVID-19 risk and intrusion were also marginally significant predictors of speed.

**Table 4.**
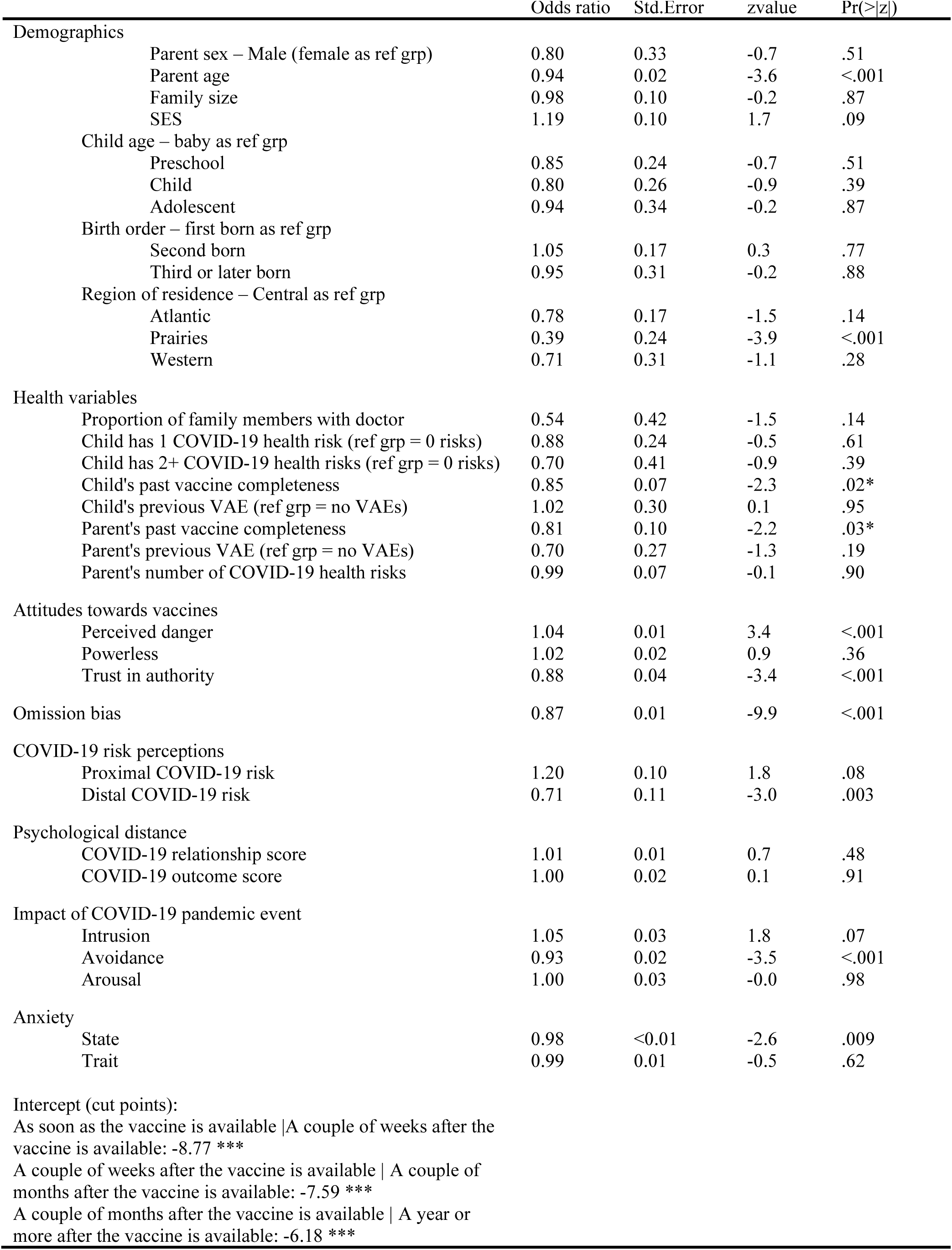
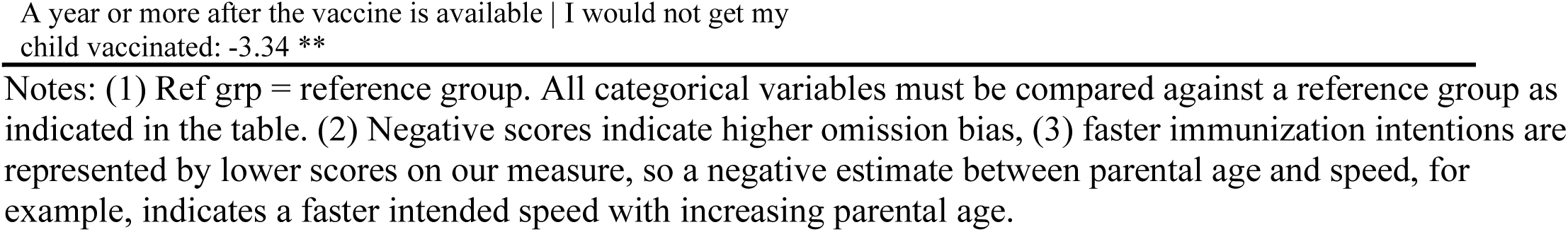
Proportional odds logistic regression model predicting speed of having children vaccinated

The polr model cannot include random effects. A Cumulative Link Mixed Model (clmm) containing both fixed and random effects in ordinal regression is therefore optimal and was thus applied. However, when including all predictor variables as in Table 4, the clmm program could not define the variance-covariance matrix, along with the standard errors and p-values, and therefore the result is not shown. The polr model was instead retained.

## Discussion

If a COVID-19 vaccine is successfully developed, vaccination of children will be required to achieve herd immunity and it is prudent to know the characteristics of those families who intend and do not intend to get their children vaccinated and how quickly this vaccination will occur. Most studies to date have only focused on vaccination likelihood, and not speed. As Canada has an especially low rate of vaccine uptake among developed countries, and has a publicly funded health care system, existing documented associations may not apply in the Canadian context. In the present exploratory study, we examined the demographic, experiential, and psychological predictors of having one’s children vaccinated.

Demographically, our results mirrored those of previous vaccine research. Parental age was positively associated with both likelihood and speed, similar to results of Taha et al. [24] regarding H1N1 vaccination intentions; Kumar et al. [21] regarding H1N1 vaccine uptake; and Niyibizi et al. [22], Galarce et al. [23], and Endrich et al. [15] regarding influenza vaccine uptake. Somewhat surprisingly, SES did not emerge as a robust predictor of likelihood, but only a trend level predictor of speed, with more affluent families indicating a faster speed of intended vaccination. This discrepancy of findings may be attributable to a number of factors including the publicly funded nature of Canada’s health care system (families would not have to pay out of pocket for the vaccine and therefore finances are not a barrier to access), and the novel context that COVID-19 has created. We are unsure of the precise reason for the increased likelihood of vaccine uptake among families residing in the Prairies, but suspect that this may have to do with political attitude differences (see e.g., ref [66]) which are associated with attitudes towards vaccination [67], and/or a desire to fully ‘re-open’ the Prairie provinces which have been especially impacted in the current pandemic (e.g., collapsing oil prices and aerospace manufacturing, [68]). For some, hopes of economic recovery hinge upon the successful creation of a vaccine [69], and so it may be that hopes for economic recovery in the Prairies hinge strongly on vaccine uptake. Future research should explore these possibilities.

Experientially, our results also mirrored that of previous vaccine research. Both parental and child previous vaccine history was associated with increased likelihood and speed consistent with the results of previous research on H1N1 [23], [29–31], seasonal influenza [32], and MMR [33] vaccination. Yet, the previous experience of VAEs did not decrease either the intended speed or likelihood of vaccination in accordance with the results of Chung et al. [36], nor did the proportion of family members with a family doctor. This is somewhat surprising given that strong relationships with primary health care providers have been associated with confidence in vaccines generally [35] and doctor recommendations are associated with H1N1 vaccine uptake [31]. However, we did not ask participants to provide detailed information on their relationships with their doctors, just whether or not they had one. It could be that simply having a doctor is not enough to promote SARS-CoV-2 vaccination intentions, but rather that the relationship between doctor and patient needs to be a strong one. Further, as the vaccine is currently unavailable, participants may not yet have discussed the possibility of a vaccine with their doctor.

Several psychologically relevant variables were associated with vaccine likelihood and speed, including attitudes towards vaccines generally. Dovetailing with previous research, those who perceived greater danger in vaccines and those with reduced levels of trust in authority relating to vaccines were less likely to endorse having their children vaccinated against SARS-CoV-2, and indicated a slower intended speed of uptake. Interestingly, feelings of powerlessness surrounding vaccination did not correlate with intended speed or likelihood of vaccinating one’s children. Powerlessness similarly did not correlate with vaccination intentions in Jolley and Douglas [39], perhaps because vaccination is largely a choice in both Canada and the United Kingdom (where Jolley et al.’s participants resided). Further, participants who preferred to accept the risks of not being vaccinated against SARS-CoV-2 (e.g., increased risk of getting COVID-19) relative to the risks of being vaccinated (e.g., side effects) therefore scored high on omission bias. Individuals high in omission bias were less likely to intend to have their child(ren) vaccinated and intended to delay vaccination for a longer period of time. This is consistent with Hamilton-West [48] who found that students high on omission bias were unlikely to receive the MMR vaccine following an outbreak of mumps on a UK university campus.

Further, psychological variables related to the ongoing pandemic were also associated with vaccine likelihood and speed. In one of our models, high levels of perceived proximal COVID-19 risk (i.e., the perceived risk of COVID-19 to oneself and one’s family) were marginally predictive of increased vaccine likelihood. This marginal association did not remain in the nested model. These results are somewhat surprising given that two existing COVID-19 vaccine intentions studies have found increased intention with increased perceived COVID-19 risk [44], [46], although, as with previous discordant results, this may be attributable to differences in decision-making processes for oneself and one’s children, as well as differences in samples. Wong et al. used a Malaysian sample and Reiter et al an American sample. American versus Canadian samples may differ in perceived levels of risk as cases have been substantially lower in Canada versus the United States [70]. Infection rates do not explain the differences between Canadian and Malaysian samples in this regard, and therefore these national differences warrant further investigation. Both America [71] and Malayasia [72] do not have universal access to healthcare, and vaccination likelihoods are probably related to this important variable.

The impact of the COVID-19 pandemic on parents, specifically on their tendency to avoid thoughts, negative emotions, or information about the pandemic, was related to an increased likelihood of having their children vaccinated, and intending to vaccinate them more quickly, whereas levels of intrusion and arousal were not related. To our knowledge no existing study has looked at these three measures in association with vaccination intentions in any pandemic context (e.g., SARS, MERS etc), and so we have little to compare our results to. However, it seems reasonable to conclude that high levels of avoidance, one symptom of post-traumatic stress disorder [60], may be associated with an increased desire to take action to potentially protect one’s family from COVID-19. Subsequent to the initiation of the present study, the Impact of Event Scale with Modifications for COVID-19 (IES-COVID19) has been developed and validated [73], and future research should include this modified scale. Levels of state anxiety were also inconsistently linked with vaccination likelihood and speed in the present study. It may also be that those who were high in state anxiety during the height of the first wave of the pandemic wish to take action to reduce risk to their families. This is partially consistent with Mohammed et al. [51] who found that mothers with mild anxiety symptoms were more likely to receive the influenza and pertussis vaccine during pregnancy than their counterparts with no or high levels of anxiety symptoms, and suggests that future research should look for curvilinear relationships between anxiety and intentions to vaccinate. In sum, higher levels of both avoidance and state anxiety were associated with increased likelihood and speed of vaccination.

Surprisingly, psychological distance from COVID-19, here measured as the number of contacts with a positive COVID-19 diagnosis, their relationship closeness, and their health outcomes was not related to likelihood or speed, contrary to Taha et al., [24] who found that participants believed their attitudes towards the H1N1 vaccine would become more positive if a member of their social circle contracted the illness. These discordant findings may be attributable to the nature of the pandemic. H1N1 was not as widespread of a pandemic as COVID-19, and many more people know someone who has been diagnosed with the disease, likely washing out some of the variability in this measure.

Overall, various demographic, experiential, and psychological predictors were related to the intended speed and likelihood of having one’s children vaccinated against SARS-CoV-2, some in ways that replicated past research on vaccination intentions, and some in surprising and novel ways. It is important that public health workers recognize the uniqueness in the Canadian context as well as the uniqueness of the family context in predicting speed and likelihood of vaccination. Understanding families’ current intentions to vaccinate for SARS-CoV-2 will allow policymakers and public health officials to develop targeted messaging campaigns to those with the greatest degree of vaccine hesitancy (see e.g., [74], [75]). Careful planning for widespread COVID-19 vaccination should begin now [2] so that, when a vaccine is developed, public health information can be disseminated in a targeted manner. This is vital research given the current prevalence of the anti-vaccination movement and how parents are promoting said movement.

Further, we recognize that the intention-behaviour gap can sometimes be large; but in at least one study intentions to vaccinate against the seasonal flu and actual behaviour were substantially correlated [76], and so we expect some degree of continuity in the attitudes parents are currently reporting. We plan to follow up with these participants to determine the predictors of vaccine uptake. Future research should include larger samples to increase power required for multi-level modelling designs, and include questions about political beliefs to further elucidate regional differences in uptake.

## Data Availability

Data are not publicly available.

## Acknowledgements

C.L. Lackner would like to thank Megan Scanlan, Mount Saint Vincent University, for her help as a research assistant throughout the project and Dave Malyk for his editorial advice. C.H. Wang would like to thank Dr. Edward Susko, Department of Mathematics and Statistics, Dalhousie University, for insightful discussions on the statistical models for the analyses.

## Notes

### Competing Interest Statement

The authors have declared no competing interest.

### Funding Statement

The research was supported by an internal Rapid Response Grant to CLL from Mount Saint Vincent University (Halifax, Canada).

### Author Declarations

Mount Saint Vincent University (Halifax, Canada) University Research Ethics Board

